# Towards automatic home-based sleep apnea estimation using deep learning

**DOI:** 10.1101/2023.02.15.23285988

**Authors:** Gabriela Retamales, Marino E. Gavidia, Ben Bausch, Arthur N. Montanari, Andreas Husch, Jorge Goncalves

**Affiliations:** Luxembourg Centre for Systems Biomedicine, University of Luxembourg, L-4367 Belvaux, Luxembourg; Department of Physics and Astronomy, Northwestern University, Evanston, IL 60208, USA; Department of Plant Sciences, Cambridge University, Cambridge CB2 3EA, United Kingdom

**Keywords:** Apnea–Hypopnea Index, Artificial intelligence, Neural networks, Automatic detection

## Abstract

Apnea and hypopnea are common sleep disorders characterized by complete or partial obstructions of the airways, respectively. A sleep study, also known as polysomnography (PSG), is typically used to compute the Apnea–Hypopnea Index (AHI), the number of times a person has apnea or certain types of hypopnea per hour of sleep. AHI is then used to diagnose the severity of the sleep disorder. Early detection and treatment of apnea can significantly reduce morbidity and mortality. However, continuous PSG monitoring is unfeasible as it is costly and uncomfortable for patients. To circumvent these issues, we propose a method, named DRIVEN, to estimate AHI at home from wearable devices and assist physicians in diagnosing the severity of apneas. DRIVEN also detects when apnea, hypopnea, periods of wakefulness occur throughout the night, facilitating easy inspection by physicians. Patients can wear a single sensor or a combination of sensors that can be easily measured at home: abdominal movement, thoracic movement, or pulse oximetry. For example, using only two sensors, DRIVEN correctly classifies 72.4% of all test patients into one of the four AHI classes, with 99.3% either correctly classified or placed one class away from the true one. This is a reasonable trade-off between the model’s performance and patient’s comfort. We use data from three sleep studies from the National Sleep Research Resource (NSRR), the largest public repository, consisting of 14,370 recordings. DRIVEN is based on a combination of deep convolutional neural networks and a light-gradient-boost machine for classification. Since DRIVEN is simple and computationally efficient, it can be implemented for automatic estimation of AHI in unsupervised long-term home monitoring systems, reducing costs to healthcare systems and improving patient care.

## Introduction

Apnea and hypopnea are common sleep-related breathing disorders that affect 6% to 17% of adults, reaching up to 49% in older populations [1]. They are usually marked by pauses in breathing or shallow breathing, respectively. This can happen, for example, when the soft tissue at the back of the throat collapses and closes at night [2]. Because blockage in the airways slows breathing, less oxygen is sent from the lungs to the heart and body. The CO_2_ level in the blood then increases due to impaired breathing, leading to episodes of sudden awakening or choking during sleep [3]. During these events, nasal airflow can cease or reduce for a short time, and the brain and body will fight to keep breathing. Sleep apnea and hypopnea have been associated with an increased risk of heart disease, diabetes, chronic kidney disease, stroke, depression, and cognitive impairment [2]. As age and the likelihood of having sleep apnea are positively correlated [4], an aging population is expected to add more pressure on healthcare systems.

Typically, the diagnosis and detection of sleep apnea and hypopnea are based on polysomnography (PSG) tests conducted in a sleep facility or at home [5]. PSG requires the overnight recording and monitoring of several physiological signals, including electroencephalogram (EEG), electrocardiogram (ECG), electrooculogram, chin muscle activity, leg movements, respiratory effort, nasal airflow, and oxygen saturation (SpO_2_). This information is then used to compute the Apnea-Hypopnea Index (AHI), the number of times a person has apnea or certain types of hypopnea per hour of sleep, as defined by the American Academy of Sleep Medicine (AASM) [6].

There are many limitations with PSG tests: they are time-consuming, expensive, and inconvenient [7]. As a result, it is expected that more than 85% of people with sleep apnea or hypopnea are not diagnosed [4]. Hence, the development of portable, easy-to-use, reliable, and affordable AHI estimation tools is crucial to improve patient care. Currently, there is only one method that estimates AHI [8], using an oximetry signal. However, this method does not estimate when a patient is sleeping, which is essential to compute AHI. Instead, it uses the information of when patients are asleep from the labels provided in the dataset (ground truth). As we will show later, oximetry signals on their own struggle to identify when patients are asleep, even when coupled with heart rate signals. This limitation can lead to poor AHI estimation. Finally, the available method does not detect events or segment oximetry signals, i.e., it does not identify the exact periods of time when apneas and hypopneas occur.

When focusing only on short time-series segments, there are many classification tools to decide whether an apnea event is present or not (see, for example, the comprehensive reviews in [9–11]). Most results are limited to obstructive sleep apnea (OSA), the most severe and common type of sleep apnea [12]. Such detection tools for OSA classification have been based on manual feature extraction, from time and/or frequency domains of one or more physiological signals. These algorithms require expert knowledge for the design and selection of features to be extracted, which may still not include the most relevant features (information) for classification from data [13]. The procedure can be hard, time consuming, and often subject to bias or lack of generalizability [14, 15]. Deep learning, on the other hand, has shifted data modeling away from “expert-driven” feature engineering toward “data-driven” feature extraction [8, 11] and is now widespread across many applications.

Based on deep learning, this paper proposes a data-driven method, named DRIVEN, to estimate AHI and segment physiological signals. We focus on devices that can be implemented in a home care environment and were available in the considered datasets, including abdominal and thoracic movement, oxygen saturation, and R-to-R interval data. Our study employs an extensive multicenter database containing 14,370 recordings of 10,752 patients belonging to multiple ethnicities and with an average age of 68.7 years. To estimate AHI, DRIVEN detects apnea and hypopnea events, and determines when patients are sleeping or awake. To our knowledge, it is the first standalone algorithm to estimate AHI. Using abdominal movement and oximetry sensors, DRIVEN correctly classifies 72.4% of all test patients into one of the four AHI classes, with 99.3% either correctly classified or placed one class away from the true one, a reasonable trade-off between the model’s performance and patient’s comfort. Cross-validation and out-of-the-domain testing show that DRIVEN is highly generalizable, achieving high performance on large external datasets not included during the training process. Given the high performance and small computational cost of DRIVEN, we expect that it could be implemented as part of a home AHI estimator system.

## Results

### Data description

Three different datasets were considered in this work: the Multi-Ethnic Study of Atherosclerosis (MESA) [16], the Men Study of Osteoporotic Fracture (MrOS) [17], and the Sleep Heart Health Research (SHHS) [18]. These datasets are publicly available from the National Sleep Research Resource for Sleep-Related Studies (NSRR) [19]. In total, 14,370 PSG recordings were considered in our study. Each PSG recording consists of data collected from several physiological sensors during a patient’s overnight sleep (lasting 8.78h on average). The PSG recordings were collected during typical PSG evaluations in healthcare facilities.

This study focuses only on channels that can be easily implemented in a home environment and are accessible in all three datasets: pulse oximetry (SpO_2_), thoracic movement, and abdominal movement. Data collected from various healthcare facilities had different sampling frequencies. These were normalized by resampling all channels to the maximum sampling frequency (64 Hz). Hence, all signals per patient were standardized to the same length, which simplified the training of the neural networks. PSG recordings were excluded from the study if they met at least one of the following criteria: (1) poor-quality PSG recordings given by the presence of “unsure” or “noise” labels in more than a third of the total sleep time or the repetition of only one value through the whole signal; and (2) the absence of one or more channels from the five initial channels selected for the study. After applying the exclusion criteria, a total of 10,643 sleep recordings were obtained to develop and evaluate the proposed method. Table 1 summarizes the databases.

**Table 1:** Data description.

| Characteristics | SHHS | MROS | MESA |
| --- | --- | --- | --- |
| Total PSG recordings | 8444 | 3871 | 2055 |
| Male | 4458 | 3871 | 953 |
| Female | 3986 | 0 | 1102 |
| Age | 64.6 (39–90) | 77.5 (67–90) | 69.4 (54–94) |
| BMI | 28.1 (18–50) | 27.1 (16–47) | 28.7 (16–56) |
| Sleep time [min] | 364.34 (34–605) | 352.19 (39–623) | 359.72 (32–601) |
| AHI [per hour] | 18.11 (0.0–161.8) | 21.57 (0.0–106.0) | 24.12 (0.0–111.9) |
| Ethnicity |  |  |  |
| White | 85.4% | 89.8% | 36.2% |
| African | 8.2% | 3.5% | 27.8% |
| Hispanic | — | 2.1% | 23.9% |
| Asian | — | 3.3% | 12.2% |
| Other | 6.4% | 1.2% | — |
| Used PSG recordings | 5288 | 3364 | 1991 |

Data are reported in mean and range (in parenthesis).
Abbreviations: Apnea-Hypopnea Index (AHI), body mass index (BMI).

The data was labeled following the definition of AHI (ahi_a0h3a) from the American Academy of Sleep Medicine [6]. An apnea is defined as a nasal flow reduction of more than 90%, while an hypopnea is defined as a nasal flow reduction of more than 30%, both for at least 10 seconds. There are several types of hypopneas, in which only the following two count towards AHI: hypopneas with a reduction of oxygen desaturation equal to 3% or more within 45 seconds (also labeled in the paper as hypopnea type 1) and hypopneas with an arousal within 5 seconds after the end of the event (hypopnea type 2) [20]. All other hypopneas (hypopnea type 3) are not considered in the calculation of AHI. In this paper, an *AHI-event* refers to any of these events: apnea, hypopnea type 1, and hypopnea type 2. According to the AASM guidelines, patients are categorized according to their AHI: healthy (less than 5 events/h), mild (5–15 events/h), moderate (15–30 events/h), and severe (more than 30 events/h).

### DRIVEN: deep learning-model for the detection of AHI-events

We developed a hybrid deep-learning model named DRIVEN to 1) classify “normal” or AHI-events, (apneas, hypopneas type 1, and hypopneas type 2) from data, 2) estimate the total time of sleep, and 3) estimate the AHI. In the classification step, the inputs of DRIVEN are short segments of time-series data recorded during sleep. We explored window lengths of 10, 30 and 80 seconds, and selected 30 seconds windows as they outperformed the others at estimating AHI (see Methods for details). We focused on the following physiological signals: SpO_2_, thoracic movement, and abdominal movement. DRIVEN then learns whether a given input corresponds to a normal event or an AHI-event. Fig. 1 illustrates DRIVEN’s pipeline. For each available physiological signal, a distinct deep convolutional neural network (CNN) is trained to extract global features from the raw 1-dimensional signals. The extracted features from all CNNs are then combined on a light gradient-boost machine (LightGBM) [21] to perform the classification between normal and AHI-event. This approach has proven to generalize better than just using one CNN to input all the channels (see subsection “End-to-end CNN” in Methods). Finally, for each patient, DRIVEN counts all detected AHI-events to estimate the AHI. We evaluate the performance of different sensors, window sizes, and deep-learning models. See Methods for details on data pre-processing and model training, validation, and testing.

**Figure 1:**
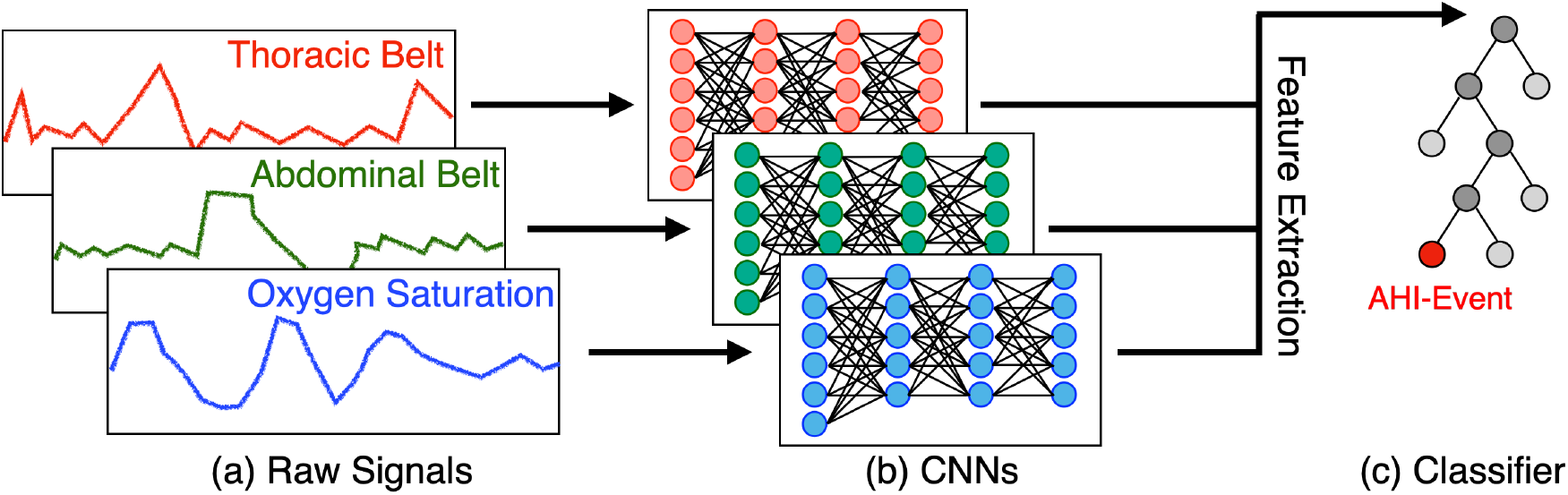
DRIVEN’s pipeline. (a) Data are separated by channels and segmented into 30 seconds windows. (b) For each channel, a distinct trained deep CNN extracts features (outputs) from the raw signal (input). (c) The extracted features are concatenated and fed to a LightGBM that classifies the input data between normal and AHI-events (apneas, hypopneas type 1, and hypopneas type 2).

DRIVEN was trained and assessed on raw physiological signals. This avoids creating human-selected features, which can sometimes lead to bias. The data were separated by centre to avoid data leakage and guarantee good performance on external datasets. SHHS was used to train and validate the model (both CNNs and LightGBM) while MESA and MROS were used as test sets to independently evaluate DRIVEN on unseen out-of-domain data from other sources. For each patient, sleep intervals were sampled every 30 seconds with a 15 seconds overlap and labeled as normal or AHI-event, yielding a total of more than 15 million samples. The training and validation data (SHHS database) were originally imbalanced with a ratio of 6.4 normal events per AHI-event. To overcome the bias and inaccuracy associated with classification using imbalanced data between two classes, the number of normal events was under sampled. Balancing the number of positive and negative samples is a common strategy followed in the literature [22].

### Performance of DRIVEN on sleep/awake detection

The AHI score is defined as the quotient between the total number of AHI events recorded overnight and the total time that patient slept (see subsection “AHI and severity class estimation from positive events” in Methods). Hence, to get an accurate estimation of this number, DRIVEN also needs to estimate the time the patient slept through the night. We trained the same architecture proposed in Fig. 1, using window sizes of 30 seconds, with the annotated labels from the dataset for awake or sleep. The training, validation, and testing methodology were similar as before. For the combination of abdominal and SpO_2_ sensors, we obtained an area under the receiver operating characteristic curve (AUROC) of 0.87 and an area under the precision-recall curve (AUPRC) of 0.90 in the test dataset. We also observe that adding RRI to any combination of sensors has little effect on the performance of the algorithm and that pulse oximetry (SpO_2_) alone is a poor sleep state predictor. The complete results of the sleep/wake classification are reported in Fig. S1 and Table S1.

### Performance of DRIVEN on classification of AHI-events

We now evaluate the performance of DRIVEN in the classification task between normal and AHI-events. Results for the complete test set (MESA and MROS databases) are shown in Figs. 2a,b using either a single channel or a combinations of channels (when the sleep/awake states are also predicted by DRIVEN). AUROC and AUPRC show that models using thoracic and abdominal movement sensors each feature very similar performance, even when these two sensors are used in combination. This indicates very high mutual information between these sensors. On the other hand, adding SpO_2_ significantly improves the performance, indicating additional information carried by this signal. Note that the model using the SpO_2_ channel alone struggles to classify AHI-events due to its poor performance to estimate sleep/awake segments (Fig. S1 shows a relatively low AUROC and AUPR of 0.72 and 0.76, respectively, on the sleep/awake detection when using only SpO_2_).

**Figure 2:**
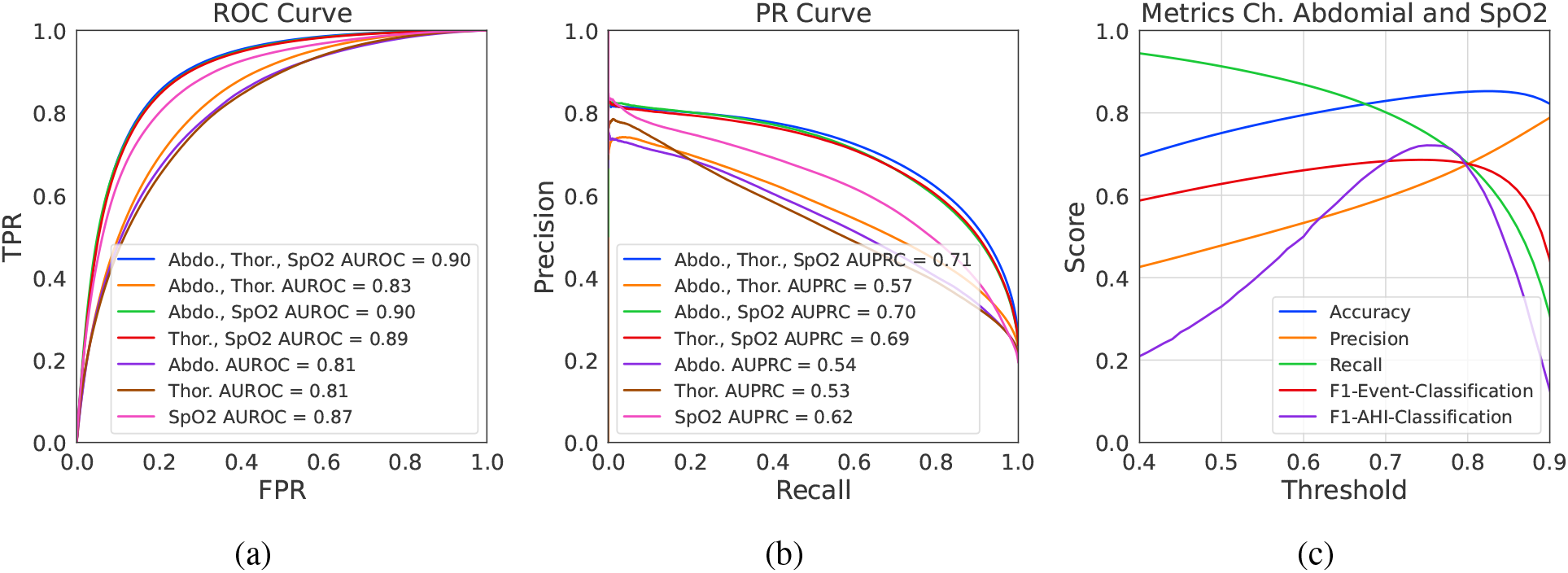
Performance of DRIVEN on classification of AHI-events when 30 seconds data segments are classified as sleep for all patients in the test dataset. (a) Receiver-operator characteristic and (b) precision-recall curves. Note the overlapping curves for thoracic and abdominal sensors. (c) Threshold dependent performance metrics of DRIVEN when using two input channels (abdominal movement and SpO_2_). The accuracy, precision, recall, and F1-Event-Classification are metrics on the classification of individual events. The F1-AHI-Classification measures the F1-Score of predicting the AHI severity category (healthy, mild, moderate, severe) on whole sleep studies.

Fig. 2c shows performance metrics as a function of the threshold when using two sensors: abdominal movement and SpO_2_. Both accuracy and F1-Event-Classification curves are relatively flat for a wide range of thresholds showing that the model is robust to the choice of threshold (which was decided on the validation results). Note that estimating hypopneas type 2 requires detecting short-term arousals, which are hard to estimate from the three available sensors. In contrast, apneas and hypopneas type 1 are much easier to detect (Fig. S2).

### Performance of DRIVEN on AHI estimation

This section evaluates the performance of DRIVEN at estimating the AHI for each patient on the test set. It consists of first estimating the total sleep time and then counting all detected AHI-events throughout the night for each patient. Using both abdominal movement and SpO_2_ sensors, Fig. 3a shows the real versus predicted AHI while Fig. 3b contains the confusion matrix of the four severity categories. In total, 72.4% of patients are correctly classified and 99.3% were either correctly classified or were placed one class away from the true one. No healthy patient was classified as severe. Among the 5,355 test patients, only *one* severe individual was classified as healthy. This occurred because the patient had a short sleep duration of 3.5 hours with frequent apneic bursts. DRIVEN exhibits consistent performance across all AHI severity levels, effectively identifying sleep-disordered breathing events in all patient groups. The model provides a good trade-off between accuracy and patient’s sleep comfort, when compared to a full polysomnography. Results for other combination of sensors are reported in Figs. S3–S8.

**Figure 3:**
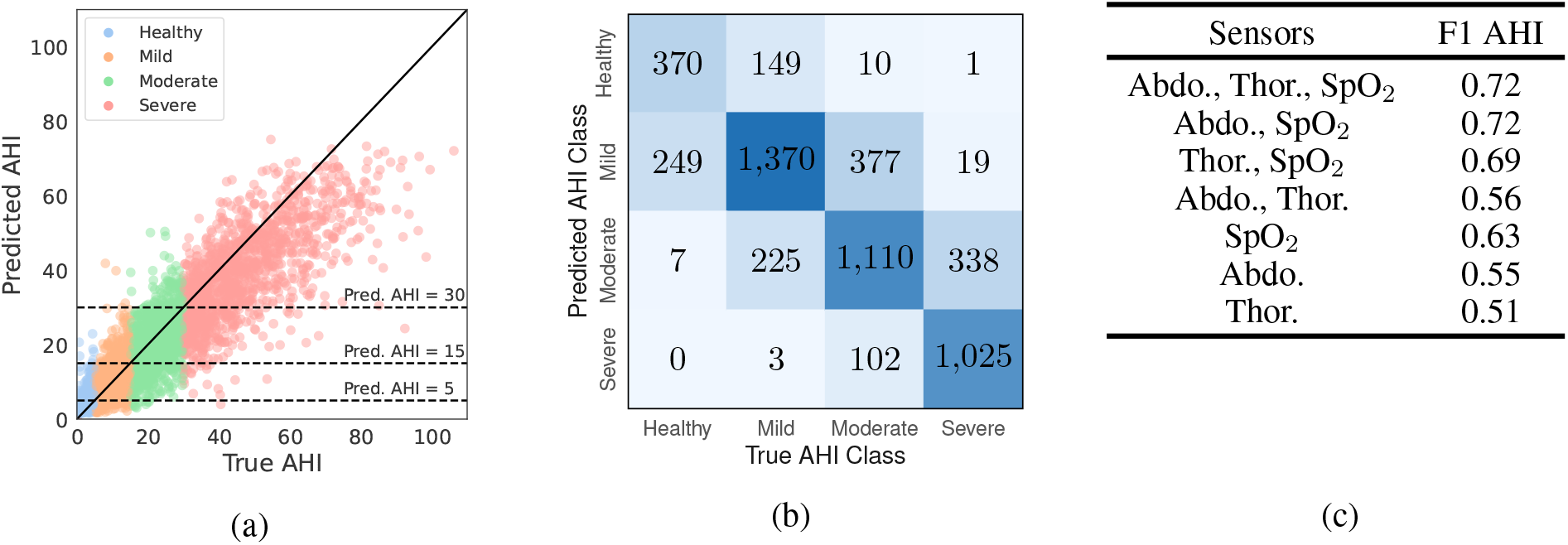
Performance of DRIVEN at estimating AHI when using two signals (abdominal movement and SpO_2_) on the test data with a threshold of 0.79. (a) Real versus predicted AHI divided by the four AHI severity groups. (b) Confusion matrix. (c) F1 scores for different combinations of sensors.

To investigate whether some physiological signals may contribute more than others in estimating AHI, we calculated F1 scores for different combinations of sensors (Fig. 3c; further details can be found in Figs. S3–S8). Similar to the classification results above, thoracic and abdominal movement sensors each have similar performance, while adding SpO_2_ significantly improves the performance. The combination of abdominal movement and SpO_2_ sensors results in one of the highest performance and using only two sensors. We also compared the AHI classification F1 scores when AHI is estimated 1) without sleep information, 2) with ground truth on the sleep/awake states, and 3) with the predicted sleep/awake states (Table S2). Using abdominal and SpO_2_ sensors, the F1 performance drops around 4% from the ground-truth sleeping information to the predicted sleep. For SpO_2_ alone, however, the drop is considerably larger at 12%, as this sensor alone cannot estimate sleep states as well.

### Automatic detection of AHI-events: segmentation of whole sleep studies

Using DRIVEN’s classification of AHI-events, we can analyse whole sleep studies to identify apnea and certain hypopneas (types 1 and 2). Additionally, with DRIVEN’s sleep state detection, we can also identify when the patient is asleep and when the patient is awake. For each patient, recordings were sampled every 15 seconds with 30 seconds windows. Each window was labeled as awake or sleep according to the sleep prediction classification. Then, those segments classified as sleep are further categorized into normal or AHI-events. Figure 4 illustrates the method on a random patient, revealing the periods of sleep with the highest AHI-event episodes, compared with the ground truth, as well as awake segments.

**Figure 4:**
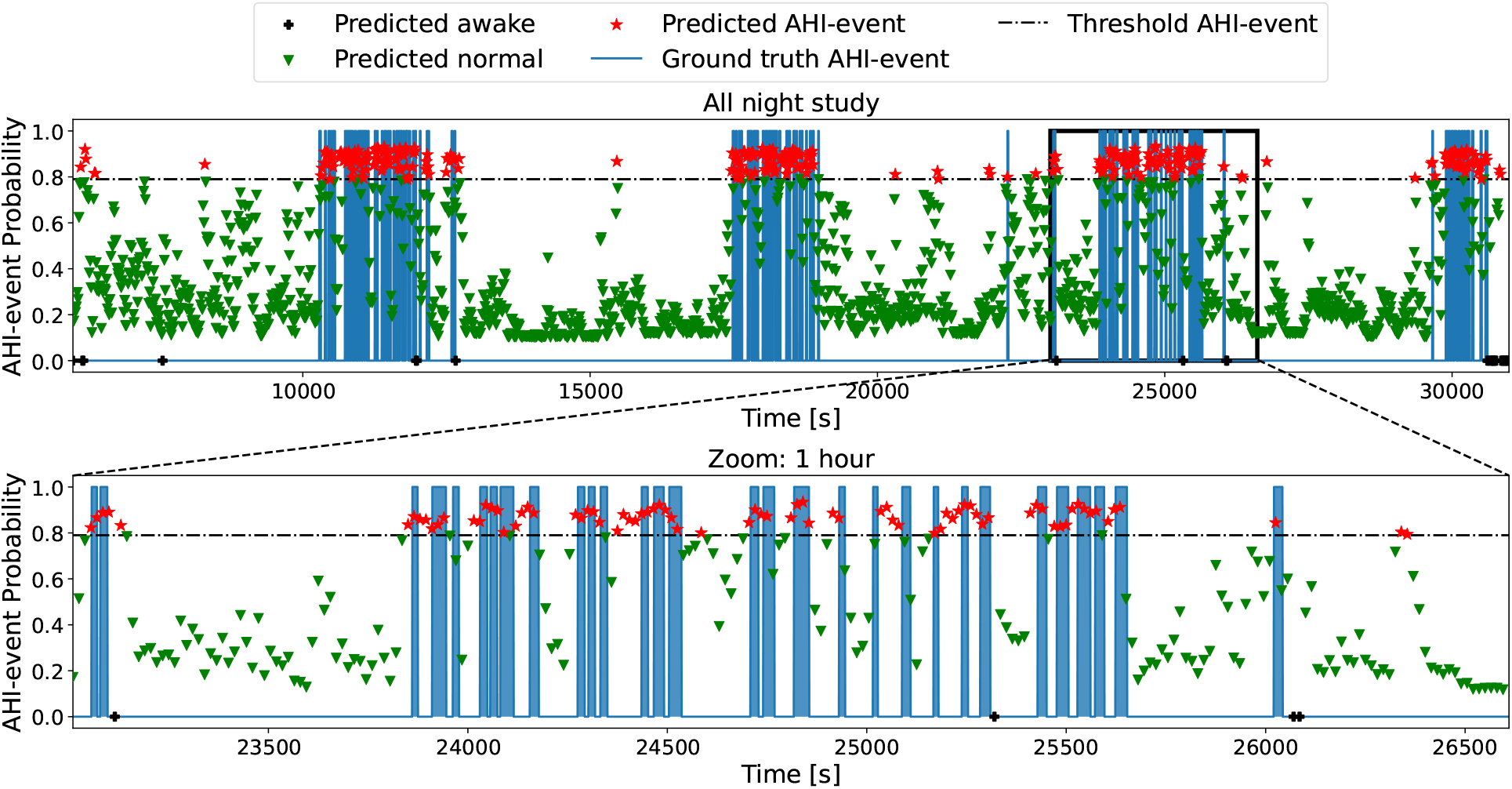
Automatic labeling of AHI-events of a random patient using two sensors (abdominal movement and SpO_2_). The blue areas represent true events (zero indicates no event and one indicates an AHI-event). The output of DRIVEN is illustrated with symbols that represent, for each 30 seconds window, the probability of the window to be classified as an AHI-event. The windows are colored according to their classification, depending on whether they are above or below the determined threshold of 0.79. The black crosses represent the segments that were classified as awake, the green triangles the ones classified as not AHI-events, and the red stars are the windows classified as AHI-events. The second plot zooms in a 1h segment. Fig. S10 increases even further the resolution to a 15 minutes interval. Fig. S11 includes the ground truth labels divided by apnea and different hypopnea types.

## Discussion

This paper introduced a method, DRIVEN, to 1) classify AHI-events and sleep states, 2) estimate AHI, and 3) segment whole sleep studies, using a single or a combination of easy to use at home sensors: abdominal movement, thoracic movement, and oxygen saturation. We showed that a deep neural network could be effectively trained using an extensive database to extract relevant features from raw physiological data. The detection was enhanced by combining it with a LightGBM classifier, which proved to generalize better than using a CNN on its own [23]. Moreover, DRIVEN’s architecture allows for each channel to have its own sampling frequency, and it estimates both AHI-events and sleep stages. The best trade-off between model performance and patient comfort was achieved when combining data from only two sensors (abdominal movement and pulse oximetry), yielding an F1-score of 72% at estimating AHI. Hence, DRIVEN can provide an estimation of AHI and detection and segmentation of whole sleep studies to aid physicians make a final diagnosis. For patients, it can guide them to seek medical attention.

Thoracic and abdominal movement sensors each featured very similar performance, since they both measure breathing. Adding SpO_2_ significantly improved the performance, which is expected given the role of SpO_2_ in the clinical definition of AHI. However, SpO_2_ alone struggled to estimate the total sleep time per patient and, for this reason, it should not be used alone. Hence, the combination of SpO_2_ and abdominal movement sensors provided an overall good performance both in detecting AHI-events and estimating the total sleep time per patient. Table S3 reports the performance of DRIVEN on AHI-event classification using all possible combinations of physiological signals available in the considered datasets. RRI data, extracted from ECGs, provided only a marginal improvement combined with other sensors. On its own, it resulted in a poor performance. Nasal airflow was also not included in the main analysis (Figs. 2 and 3) since it is an uncomfortable sensor to sleep with, and provided similar performance than either thoracic movement or abdominal movement sensors.

The reasons for choosing CNNs to automatically search for features over domain knowledge feature extraction methods are mainly due to the variety of channels to evaluate and the large available datasets. The flexibility of deep-learning feature extraction approach allowed us to explore several choices of physiological channels (including all possible combinations). DRIVEN’s architecture allows us to easily and efficiently evaluate new sensors, such as adding a movement sensor if it would be available. Several studies have shown that, when the training datasets are large and there is enough computing power, deep learning performs as well or better than other machine-learning algorithms [24], as demonstrated in different applications ranging from chess games and speech recognition to computer vision [25]. In the particular case of obstructive apnea detection, Ref. [26] reported consistently better results using unbiased deep learning approaches and CNNs for segment classification in PSG studies.

A general limitation, not just of DRIVEN but also any other method without access to EEG data, is detecting hypopneas type 2. This type of hypopnea requires estimating short-term arousal following a drop of at least 30% in nasal airflow, which is particularly challenging to detect from the three sensors selected in this study (Fig. S2). Figures 4 and S10 also show a few false positives and some 30 seconds segments with a high probability of being AHI-events, which in fact are hypopneas type 3 (that are not considered in AHI). Indeed, detecting AHI-events (apneas and hypopneas types 1 and 2) is considerably more challenging than simply detecting obstructive sleep apneas (Fig. S2), which is the main focus of most available algorithms [26]. A more specific limitation of DRIVEN is its use of 30 seconds windows, which might treat bursts of multiple short-lived apnea or hypopnea as a single long event. While the smoothing window has merits by effectively filtering noise and capturing longer events, it could lead to underestimating the number of apnea or hypopnea, especially in cases of severe AHI. This effect can be seen in Fig. S10, which is a zoomed-in segment of Fig. 4. The smoothing effect caused individual apnea events of bursts to be merged into one event, resulting in a lower AHI estimation. The complex influence of different bursting patterns of apnea events on AHI is also discussed in the literature. For example, Ref. [27] demonstrated that very different bursting patterns could lead to the same AHI values. In future studies, we plan to explore new types of AI models, including selective state-space neural networks [28], to potentially further improve performance and address these limitations. Finally, we investigated classifying sleep or awake from combinations of the three sensors considered in the paper (Table S1 and Fig. S1). There was a decrease in performance of DRIVEN when using predicted sleep (Figs. 2) instead of the annotated total sleep time per patient obtained from the dataset (Fig. S12), as summarised in Table S2. In practice, DRIVEN’s performance could be further improved by combining these sensors with other easy to wear sensors like smartwatches or sensors under the mattress.

This result demonstrates the potential of DRIVEN for an automatic labeling application. Compared to a Polysomnographic Technologist’s manual detection and diagnosis of OSA, computer-assisted signal analysis systems can reduce errors related to inter- and intra-operation variability and tiredness induced by the arduous annotating process [29]. Furthermore, most computer-based analyses can be conducted more cost-effectively and quickly [13]. Overall, DRIVEN could reduce the burden on Polysomnographic Technologists and costs to the healthcare system by making feasible the examination of patients through multiple nights from data that can be collected at home by wearable devices.

## Methods

### Data specification

The polysomnography data is available in European Data Format (EDF) [30], while the profusion files, in XML format, serve as the annotated label files. These files have been annotated for each sleep stage using the Rechtschaffen & Kales (R&K) criteria [31]. Each profusion file includes annotations for every 30-second sample of sleep stages and instances of sleep problems, with information of initiation and duration time, as well as the sensor used to detect the anomaly. Databases are summarized as follows.

1. The MrOS Sleep Study: MrOS is a substudy of the Men Study of Osteoporotic Fractures. Between 2000 and 2002, a baseline evaluation was performed on 5994 elderly men 65 years of age or older taken from six clinical institutions. Between December 2003 and March 2005, a total of 3,135 of these participants were subjected to complete unattended polysomnography and 3 to 5-day actigraphy tests as part of the sleep study. The purpose of the Sleep Study was to determine the extent to which sleep disorders are linked to adverse health outcomes, such as the increased risk of death, fractures, falls, and cardiovascular disease.
2. The MESA Sleep Study: The Multi-Ethnic Study of Atherosclerosis (MESA) is a collaborative 6-center longitudinal investigation of factors associated with the development and progression of subclinical to clinical cardiovascular disease in 6,814 black, white, Hispanic, and Chinese American men and women aged 45 to 84 years in 2000-2002. The participants were all between the ages of 45 and 84 at the time of the study. Four follow-up examinations have been performed, one each in the years 2003–2004, 2004–2005, 2005–2007, and 2010–2011. Furthermore, 2237 people participated in a Sleep Exam conducted by MESA Sleep between 2010 and 2012. This exam included an unattended overnight polysomnogram, wrist-worn actigraphy for seven days, and a sleep questionnaire. The sleep study aims to determine whether there is a correlation between subclinical atherosclerosis and sex, ethnicity, or other demographic differences in sleep and sleep disorders.
3. The SHHS Sleep Study: The Sleep Heart Health Study is a multicenter cohort study conducted by the National Institute of Heart, Lung, and Blood. The purpose of the study was to investigate the effects of sleep-disordered breathing on the cardiovascular system and also on other aspects of a person’s health and to determine whether breathing problems that occur during sleep are related to an increased risk of coronary heart disease, stroke, death from all causes, and hypertension. Between November 1995, and January 1998, SHHS Visit 1 research focused on participants who were at least 40 years old and included 6,441 men and women. A second polysomnogram, known as SHHS Visit 2, was performed on 3,295 participants during the third exam cycle (January 2001 to June 2003).

For the labelling of events, a window is considered AHI-event positive if it is satisfies one of the AHI conditions (apnea, hypopnea type 1 and hypopnea type 2). To obtain AHI, the total number of AHI-events throughout the night is divided by the total number of hours of sleep.

### Data pre-processing and model training

First, data is split by measurement channel (SpO_2_, abdominal movement, thoracic movement, RRI and airflow). For each patient, the initial and final 30 minutes were removed from the recording as they are considered set up times. Then, for each channel per patient, all signals are standardized and normalized (to speed up training) by calculating the z-scores of the 95% of the data points and applying min-max normalization to eliminate the bias of mean and variance of the raw one-dimensional signal [32]. The RRI data was inferred from ECG data by measuring the time difference between heartbeats (from one R peak to the next R peak) using the Pan-Tompkins algorithm [33, 34]. Subsequently, each channel’s frequency was set to 64Hz, which is the maximum frequency of the measurements, using the cubic spline as the interpolation method [35]; this method is simple to implement and, at the same time, it does not attenuate higher frequency components of the signal. Finally, the signals were segmented into windows of 30 seconds and labeled as “normal” and “AHI-events”. Data windows correspond to the same time instance across all channels (Fig. 1a).

Normalized samples of 30s from each channel are then fed in parallel into distinct deep CNNs. Each CNN is independently trained on a specific channel to classify the AHI-events from the normal windows, and then used for automatic feature extraction (Fig. 1b). The training was performed balancing the number of normal and AHI-events fed to the network. For this task, the number of normal samples was randomly downsampled to match the amount of positive samples. Once the networks have been trained, features are extracted and concatenated to integrate information from different physiological biomarkers. This is achieved by using the concatenated features to train a LightGBM classifier for binary classification between normal and positive AHI-event windows (Fig. 1c).

The CNNs were trained and cross-validated with 7,753,500 samples, 1,042,652 positives and 6,710,848 negatives, from 5,288 PSG recordings in the SHHS database. We use the EfficientNetV2 architecture, a deep CNN with 479 layers developed by Google in 2021 [36]. It is a modified and optimized version of EfficientNet [37], a popular image classification algorithm that won the ImageNet 2019 competition [38]. The architecture used in this paper has been modified to handle unidimensional data and perform binary classification. Each CNN was independently trained using raw unidimensional physiological data from pulse oximetry, RRI, thoracic movement, abdominal movement, or nasal airflow. Categorical cross-entropy was used as the loss function and ADAM as the optimizer [39]. If the validation loss did not decrease after eight consecutive epochs, the training was terminated. Once the networks were trained, the final two layers were removed and the last global average pooling layer is used to yield 1,280 features from each data channel.

### Feature classifier

After the neural networks are trained, and features are extracted, the next step is to concatenate the features extracted by all CNNs and use them for training a LightGBM classifier. In contrast to a large number of other well-known algorithms, such as XGBoost [40] and GBDT [41], LightGBM employs the classification algorithm for growing trees in a leaf-wise manner rather than in a depth-wise manner. The leaf-wise algorithm can converge significantly faster than the depth-wise growth method, although its growth can be subject to overfitting if the appropriate hyperparameters are not used [21]. We use a random search method within a specified set of parameters to optimize the training and performance of the LightGBM. This allows a fixed number of parameters from a particular distribution to be sampled instead of testing all the values of potential parameters [42]. The output of the feature classifier is the probability that the evaluated segment contains AHI-events according to the AHI definition. The training was performed with a balanced dataset. The complete datasets, however, are imbalanced (more negative AHI-events than positive ones). Hence, on the validation data, we determined the probability threshold that optimizes the performance metrics of the classifier. This probability threshold was chosen so that the precision and the recall metrics have the same value in the validation dataset (Fig. S9a).

### Models and channels comparison

We evaluated longer and shorter window sizes for event classification and AHI estimation on a subset of 1000 randomly selected patients in a 3-fold cross-validation experiment, and selected 30 seconds windows for their higher performance in the validation set. Additionally, we noticed a delay in the response of SpO_2_ signal when a respiratory event occurred; thus, we also evaluated the model performance using different delays in the SpO_2_ signal (10, 15, 20, and 25 seconds delays). For AHI-event classification, we evaluated the feature extraction at different layers of the neural network (model called LGBM-1 for the last layer and LGBM-3 for the third last layer), and the performance of neural networks trained on each channel separately. In summary, we evaluated 9 different channels (abdominal, thoracic, nasal airflow, RRI, SpO_2_, SpO_2_+10s delay, SpO_2_+15s delay, SpO_2_+20s delay, and SpO_2_+25s delay), two different models (LGBM applied to the combination of different sensors extracting the information at the third last layer or at the last layer of each EfficientNetV2), and three different window sizes (10s, 30s, and 80s). Note that every time SpO_2_ is used, all its delayed signals are also used as separate channels. The summary of these results are reported in Table S3 and the complete set of results is in Table S4. These tables show that the 10s windows yield consistently lower performance and RRI signals provide negligible performance improvements. Hence, we now focus on the window sizes of 30s and 80s, as well as the channels abdominal, thoracic, and SpO_2_, and use the training data as described below. The complete performance results are reported in Table S5, which also includes the estimation of AHI. On the validation dataset, these tables show that a larger window size leads to better classification of AHI-events whereas a lower window size results in better AHI estimation. Since most AHI-events last 30 seconds or less, the use of shorter windows may yield more accurate AHI estimations despite having slightly lower classification performance. Hence, we finally selected the 30 seconds windows because the objective of this paper is on AHI estimation. The 30 seconds windows also allow us to do a better whole sleep study segmentation.

### End-to-end CNN

DRIVEN’s architecture consists of a combination of CNNs and LightGBM. Here, we compare its performance with that of a single CNN that inputs all channels and then performs the final classification. For this model, we used the same NN, EfficientNetV2, and changed the input layer to accept a vector for each channel. As in the previous section, we compared the AHI-event classification as well as the AHI classification of patients with the same 1,000 randomly selected patients. Table S4 shows that the validation results are quite similar, with the end-to-end CNNs having a slightly lower performance compared with the combination of CNNs and LightGBM. However, the performance of the end-to-end CNNs drop significantly in the test datasets, which are also detailed in this spreadsheet. The end-to-end CNNs tend to overfit the training data and the combination of CNNs and LightGBM shows better generalization across datasets. This result has also been observed by others (see, for example, [23]).

### Dataset partition into train, validation, and test

We performed three different trials with the complete dataset:

1. SHHS and MROS patients as training and validation sets in a 80-20% random split and MESA patients as test set,
2. SHHS patients as training and validation sets in a 80-20% random split and MESA and MROS as test set, and
3. SHHS, MROS and MESA patients as training, validation and test sets in a 60-20-20% random split. Because there are patients in SHHS and MROS databases that have two recordings, to avoid data leakage we were careful to always place them in the same partition. The performance metrics for validation and test sets of each trial can be found in Table S5. All the results and figures presented in the paper are from trial 2, as we only trained and validated with patients from the SHHS database and tested the model’s performance to close the domain gap when applying our algorithm to the other two independent databases.

### Sleep/awake classification

The calculation of AHI requires the total time the patient has slept, excluding awake times throughout the night. Hence, it is important to discriminate between sleep and awake intervals. The profusion files per patient were already labeled with this information, which we could use to compute the total sleeping time. This information can also be estimated from other sensors like photoplethysmograms (PPG) [43] or accelerometers. Here, to provide a standalone solution, we investigated classifying sleep or awake from combinations of only the available three sensors considered in the paper. We used the same architecture (CNN+LGBM) and model comparison methodologies to estimate if a 30 seconds window is sleep or awake. Additionally, we investigated adding an extra sensor, RRI, already discarded for AHI-events prediction. The results are presented in Table S1 and Fig. S1. The performance is high for most combinations of sensors. The exception is SpO_2_ alone, which struggles to correctly differentiate between sleep and awake windows. Adding an RRI sensor does not improve the performance when the best classifiers and alone it is not a well predictor of sleep or awake. This sleep prediction, with a threshold of 0.5 to classify sleep or awake, is then used for the AHI estimation of each patient.

### AHI and severity class estimation from positive events

As the data were imbalanced, we determined the final classification threshold as the point of intersection of the precision and the recall curves on the validation data (Fig. S9a). Then, for each patient in the validation dataset, we went through the overnight study classifying all 30 seconds windows sampled every 15 seconds (hence, consecutive windows overlap by 15 seconds). Each 30s window was first classified as awake or sleep, and the sleep segments were further classified into positive AHI-event or negative by using the previously defined threshold (Fig. 4). Next, we calculated the ratio between the number of positive AHI-events over the total sleep segments. With these ratios, one per patient, we fitted a linear regression to the value of AHI of these patients. In this regression, we used weights to account for the imbalance of the number of patients in different AHI-classes. For the test set, we used the threshold selected before (Fig. S9b), and obtained the AHI-event vs sleep windows ratio as before. Finally, we used the previously determined linear regression factors to obtain the AHI of the test patients. Recall that the patient severity class was determined following the rules provided by the American Academy of Sleep Medicine [6].

### Performance metrics

The performance of the models was evaluated based on the following metrics. Let true positive (TP) represent the number of AHI-events that are accurately predicted. True negative (TN) represents the number of normal events that are accurately predicted. False negative (FN) represents the number of AHI-events incorrectly predicted as normal events, and false positive (FP) represents the number of normal events incorrectly predicted as AHI-events. The accuracy of the model is given by (TP + TN)*/*(TP + TN + FP + FN), indicating the probability of correctly identifying normal and AHI-events; the recall is given by TP*/*(TP + FN), indicating the probability of identifying AHI-events; the specificity is given by TN*/*(TN + FP), indicating the probability of detecting normal events; the precision is given by TP*/*(TP + FP), indicating the ratio between detected AHI-events and all predicted AHI-events cases; and the F1-score 2(precision *×* recall)*/*(precision + recall), indicating the harmonic mean of precision and recall. For every method, the receiver operating characteristic curve, and the precision-recall curve were obtained for each possible classification threshold. The area under both of this curves allows us to compare the global performance of each architecture. Finally, the patient severity classification results were evaluated by assessing the F1 score of each class and then averaging them.

## Acknowledgements

The authors acknowledge support from the Luxembourg National Research Fund (FNR) through grants PRIDE15/10907093/CriTiCS, AFR/17022833 and INTER/DFG/21/15020234, the latter is co-funded by the Deutsche Forschungsgemeinschaft (DFG) project number 458610525. High performance computing experiments for model training and evaluation presented in this report were carried out using the HPC facilities of the University of Luxembourg [44].

## Contributors

J.G. conceptualized the research; M.E.G., J.G., G.R., and A.N.M. developed the AI model; G.R., M.E.G. and B.B. implemented the codes and validated the results; G.R., M.E.G., J.G., B.B., A.H., and A.N.M. analyzed the results; J.G. and A.H. supervised the work; all authors wrote and revised the manuscript.

## Data availability

The data was provided by the National Sleep Research Resource and are publicly available on request at https://sleepdata.org/.

## Code availability

Data pre-processing and segmentation were implemented using MATLAB software. The neural network was implemented on the Keras framework with Tensorflow backend on Python 3.7. The codes are available in GitHub at https://github.com/LCSB-SCG/DRIVEN/.

## Supplementary Material

**Figure S1:**
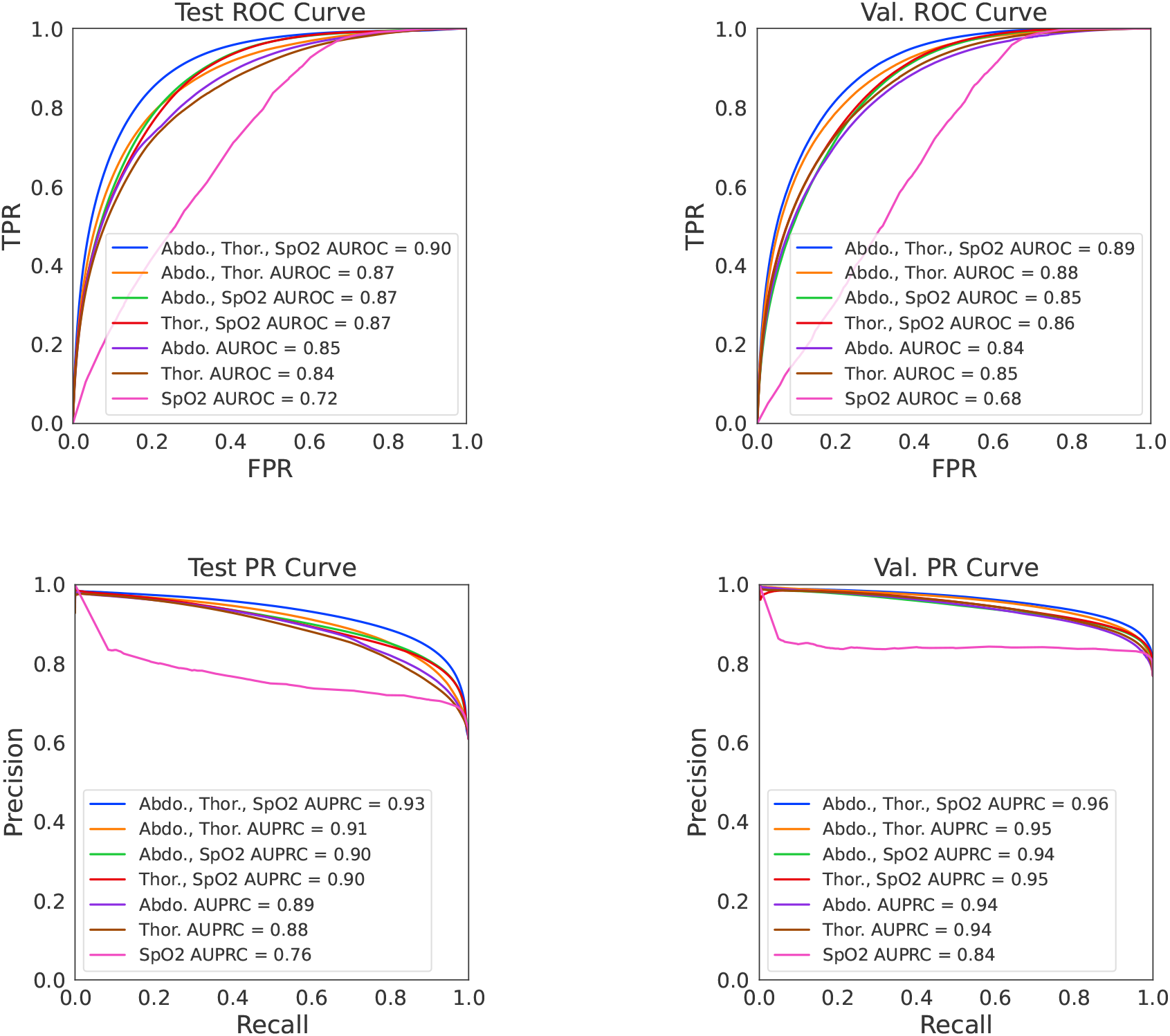
Comparison between different sensor inputs to DRIVEN for sleep/awake detection on the test (left) and validation (right) datasets. The top and bottom panels show the ROC and PR curves, respectively.

**Figure S2:**
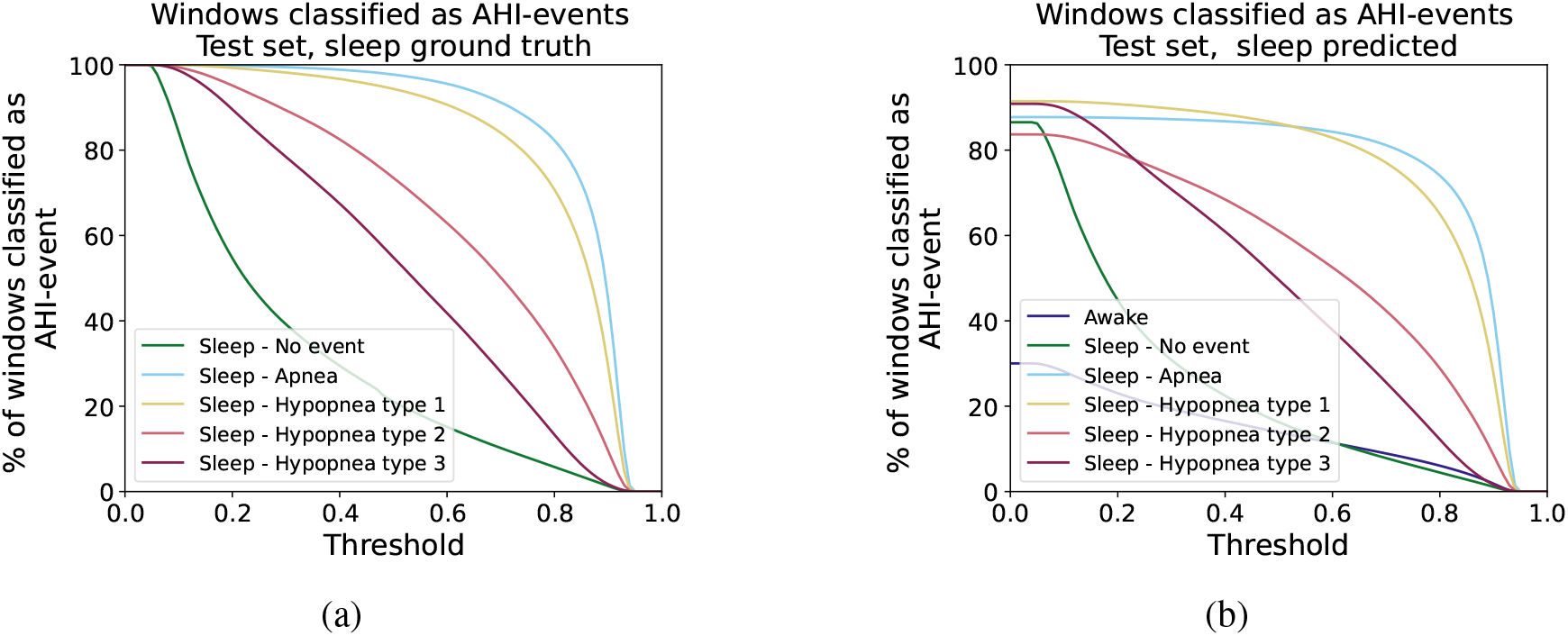
Percentage of 30 seconds windows classified as AHI-event per class on the test dataset. Each window is labeled from the profusion file as the following classes: awake, sleep without respiratory event, sleep with apnea, sleep with hypopnea type 1, sleep with hypopnea type 2, or sleep with hypopnea type 3. Each curve represents, as a function of the classification threshold, the percentage of events in the corresponding class classified as an AHI-event. The classification was performed using two input channels (abdominal movement and SpO_2_). (a) Performance results are shown when the ground truth for sleep/awake events are used (therefore no “awake” is classified as an “AHI-event”). (b) Performance results are shown when sleep/awake detection is used (therefore some “awake” windows are classified as “sleep, no AHI-event” and some are classified as an “AHI-event”). In this case, certain “AHI-events” may be misclassified as “awake” events.

**Figure S3:**
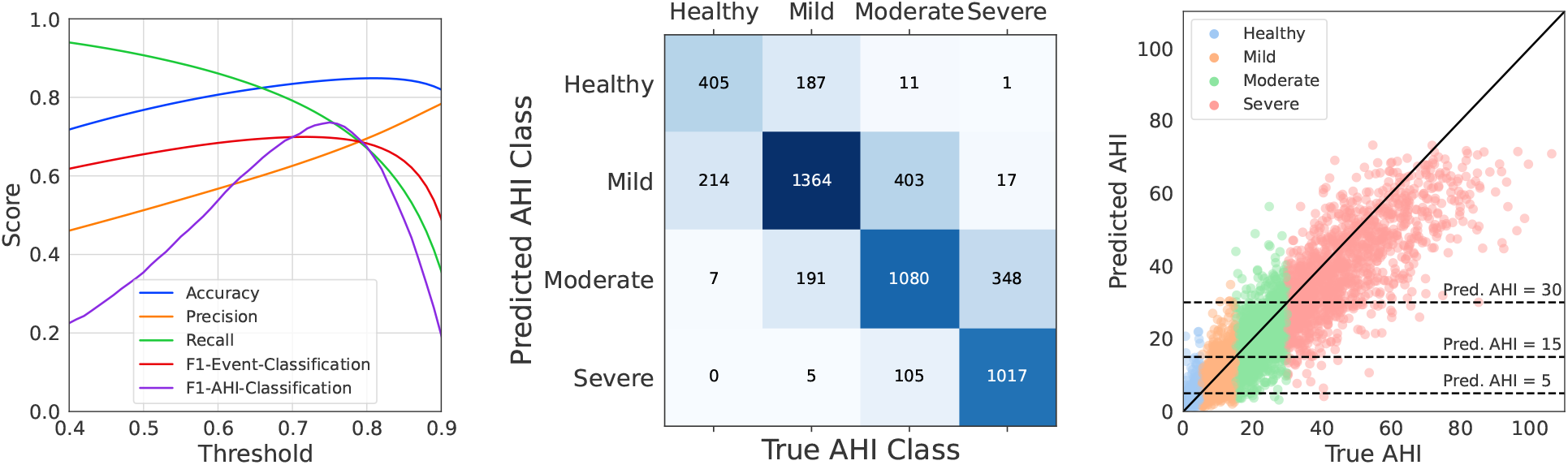
Performance of DRIVEN on AHI estimation when using abdominal, thoracic, and SpO_2_ sensors.

**Figure S4:**
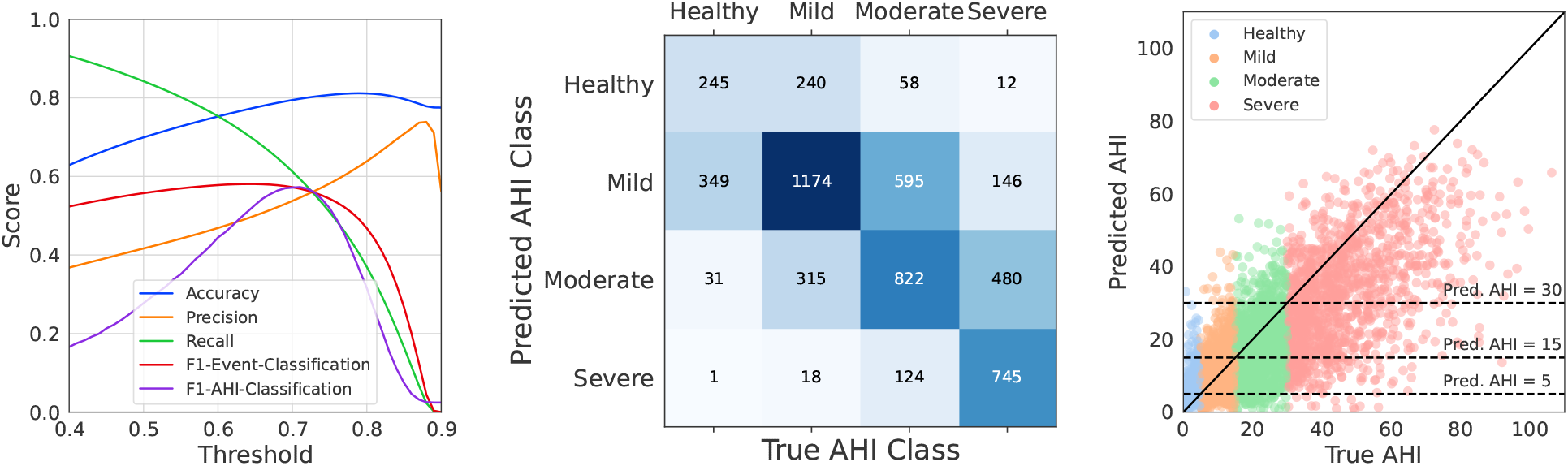
Performance of DRIVEN on AHI estimation when using abdominal and thoracic sensors.

**Figure S5:**
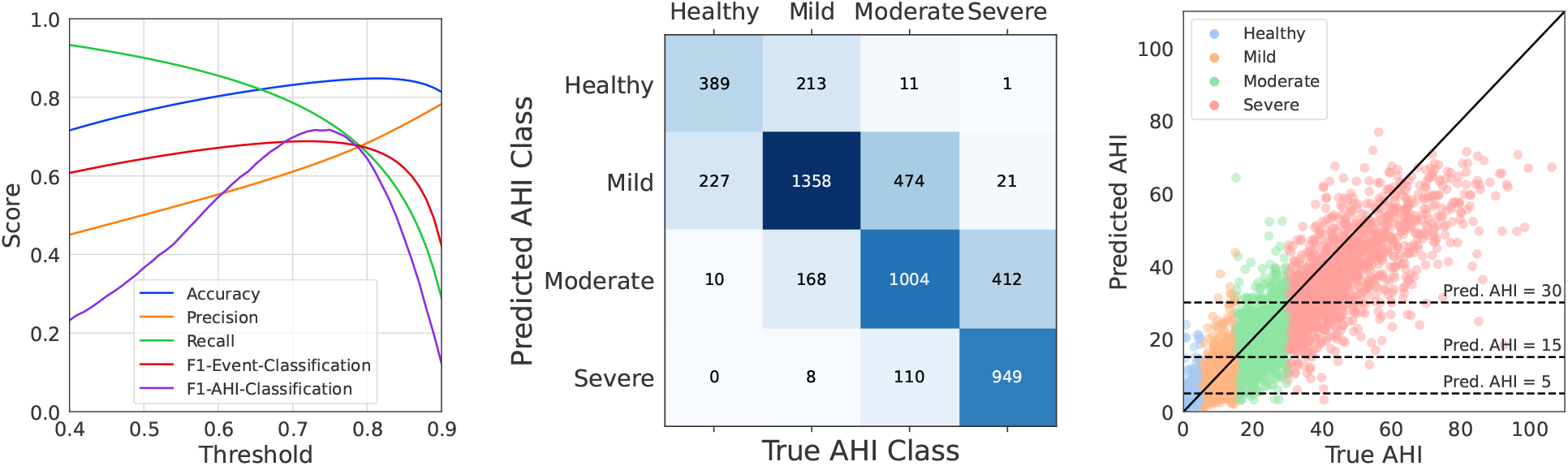
Performance of DRIVEN on AHI estimation when using thoracic and SpO_2_ sensors.

**Figure S6:**
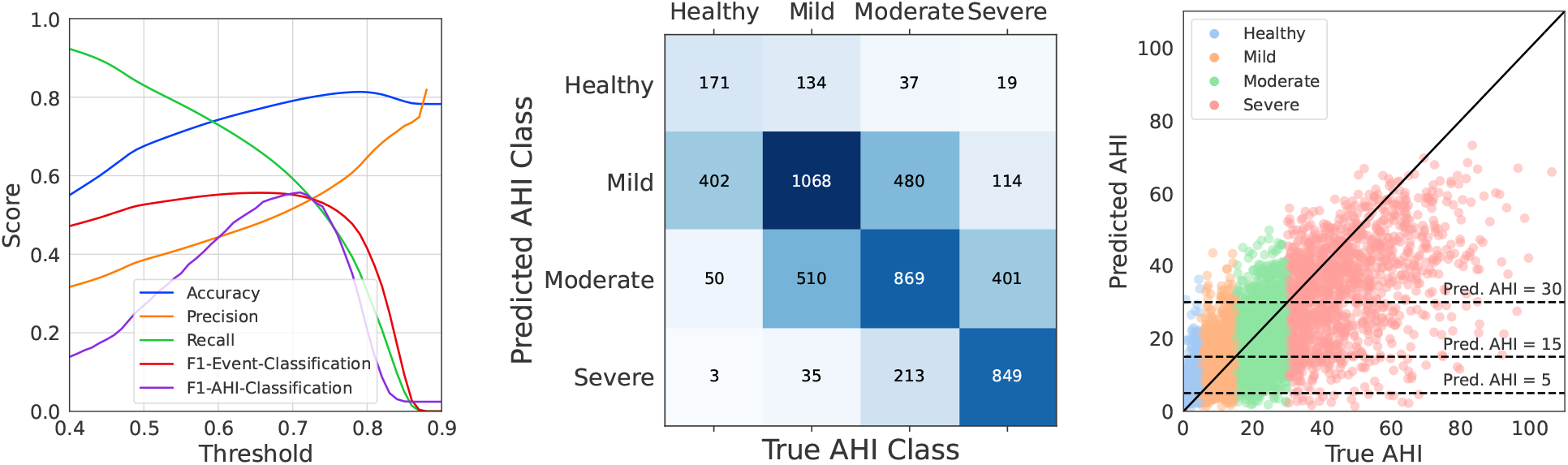
Performance of DRIVEN on AHI estimation when using only the abdominal sensor.

**Figure S7:**
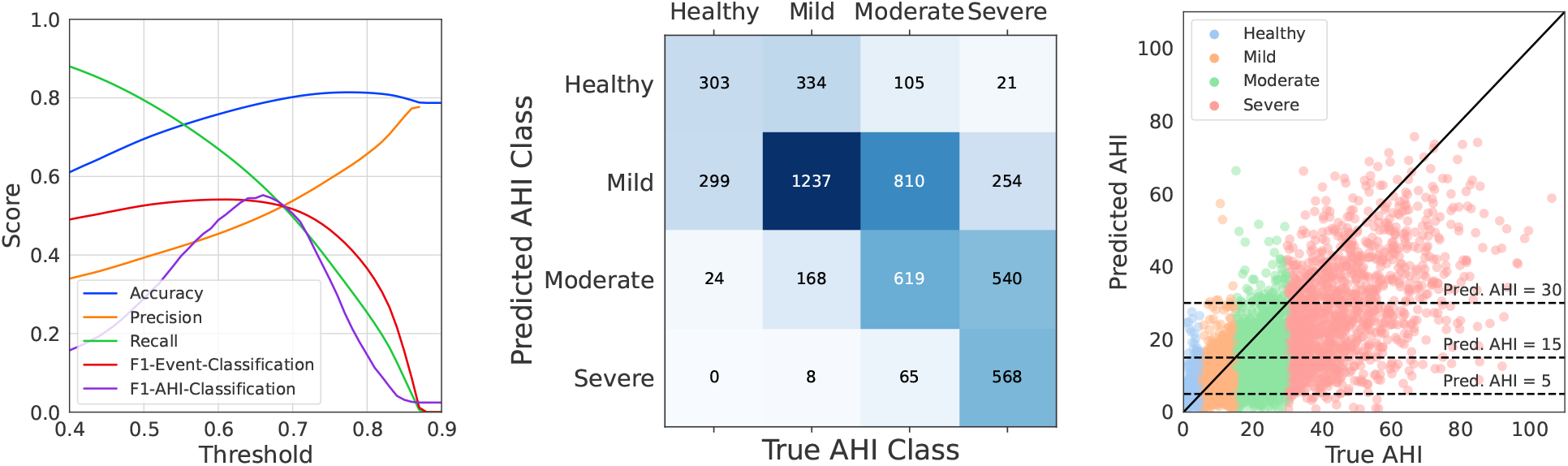
Performance of DRIVEN on AHI estimation when using only the thoracic sensor.

**Figure S8:**
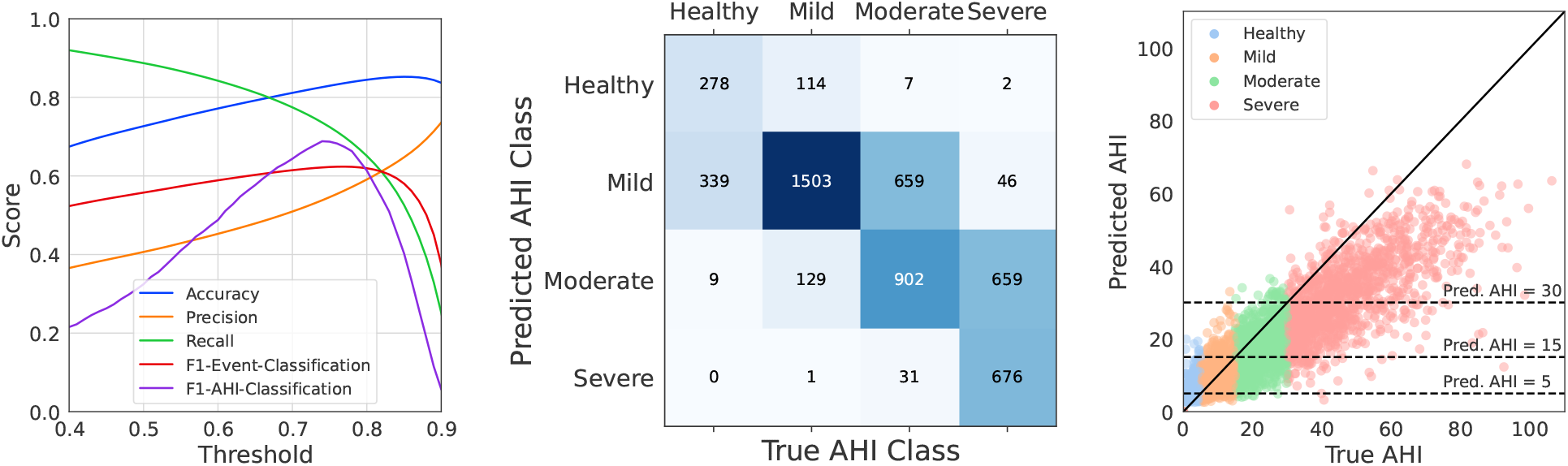
Performance of DRIVEN on AHI estimation when using only the SpO_2_ sensor.

**Figure S9:**
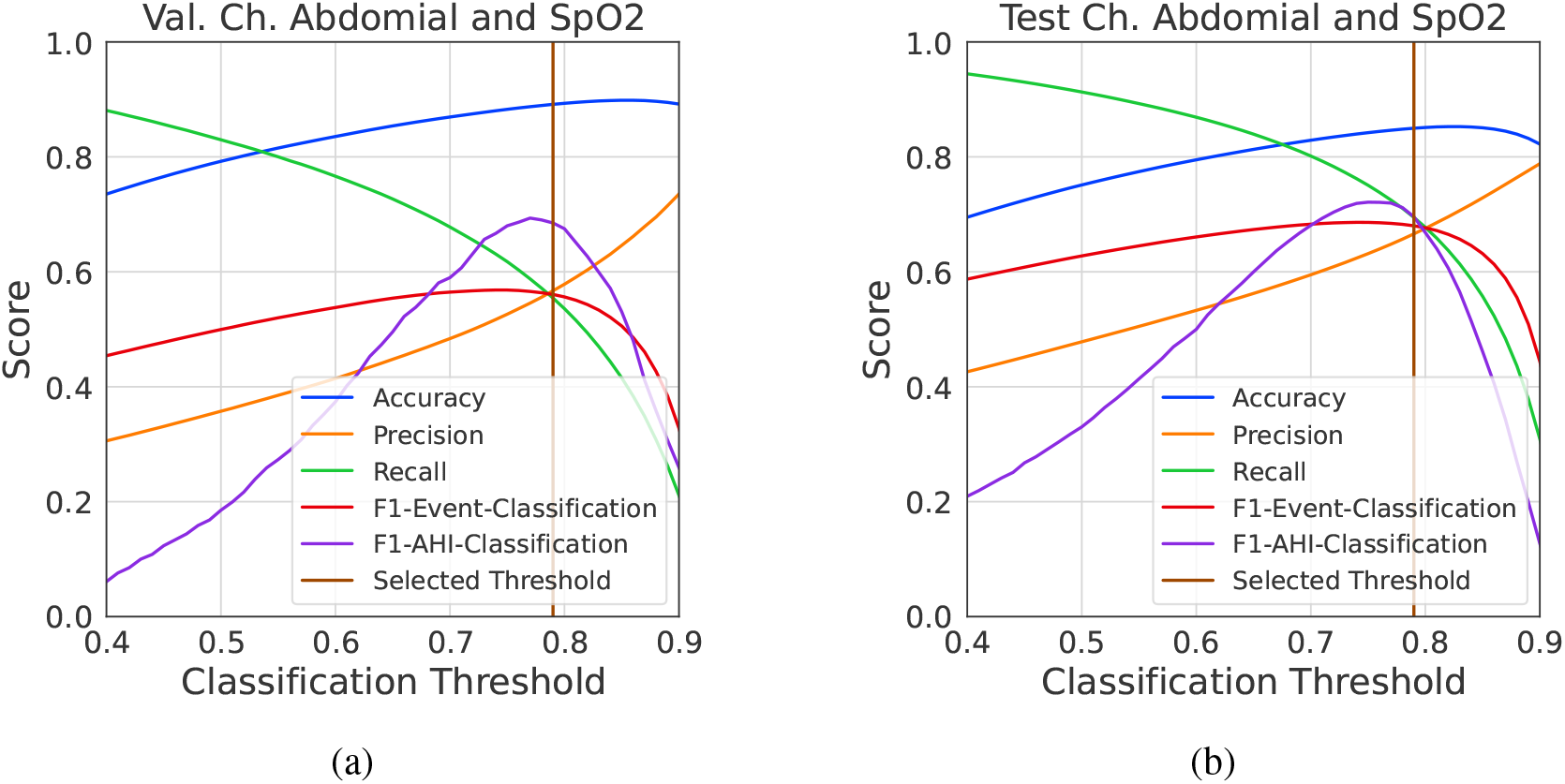
Choice of classification threshold. (a) The threshold used for classification was chosen as the point of intersection of the precision and recall curves on the *validation* dataset of trial 2. (b) Performance metrics for the *test* set, with the brown vertical line depicting the threshold chosen in panel (a).

**Figure S10:**
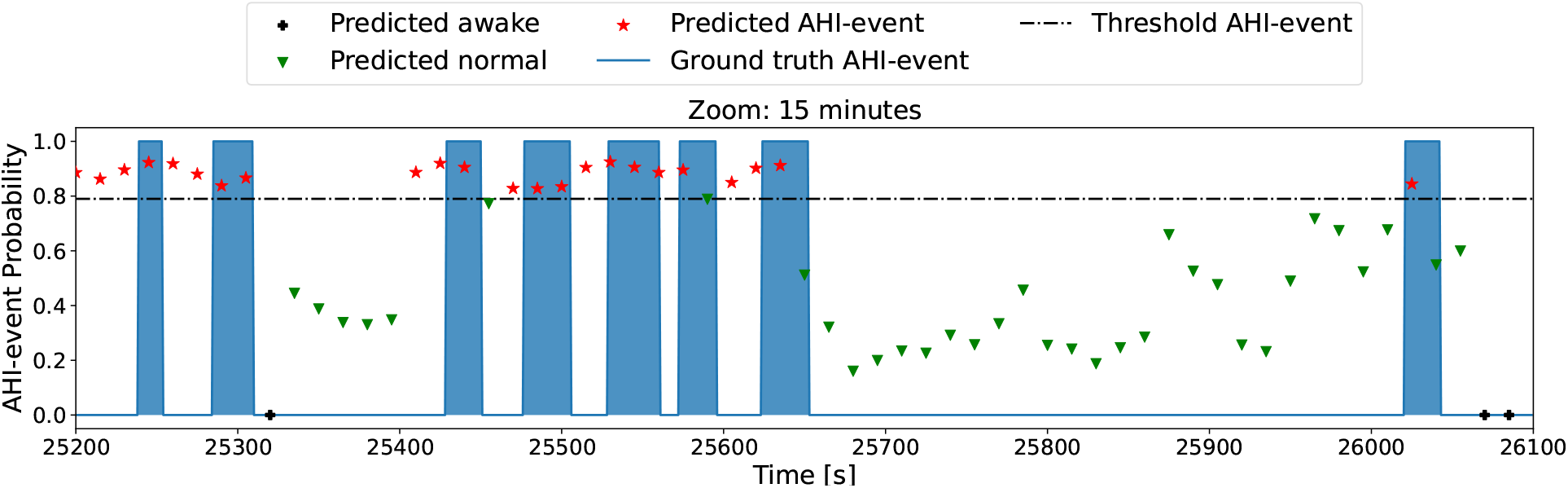
Additional 15 minute zoom-in on a section of the data shown in Fig. 4. It can be observed that detection is smoothed in-between AHI-events, which results in incorrectly detecting AHI-events between some individual events that closely follow each-other.

**Figure S11:**
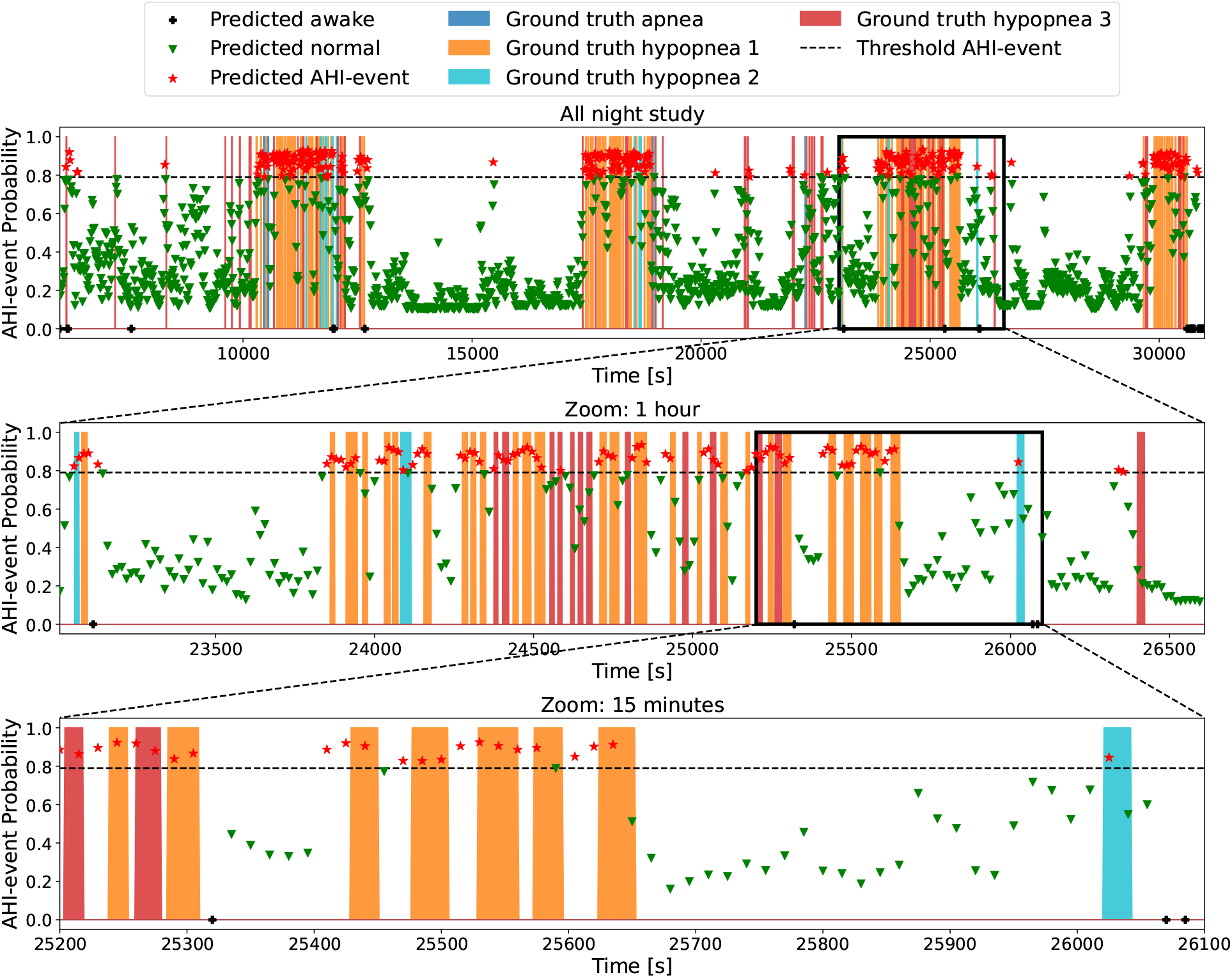
Automatic labeling of AHI-events for the same patient as Figures 4 and S10 using two sensors (abdominal movement and SpO_2_). The colored areas represent true events: blue for apnea events (flow reduction of more than 90%), orange for hypopnea type 1 (flow reduction of more than 30% and SpO_2_ desaturation of more than 3%), cyan for hypopnea type 2 (flow reduction of more than 30% and arousal), and red for hypopnea type 3 (not considered in AHI). The output of DRIVEN is illustrated with symbols that represent, for each 30 second window, the probability of the window to be classified as an AHI-event. Finally, the windows are colored according to their classification, depending on whether they are above or below the determined threshold of 0.79. The black crosses represent the segments that were classified as awake, the green triangles the ones classified as not AHI-events, and the red stars are the windows classified as AHI-events. Going down, the second and third plots zoom in a segment of 1h and 15 minutes, respectively.

**Figure S12:**
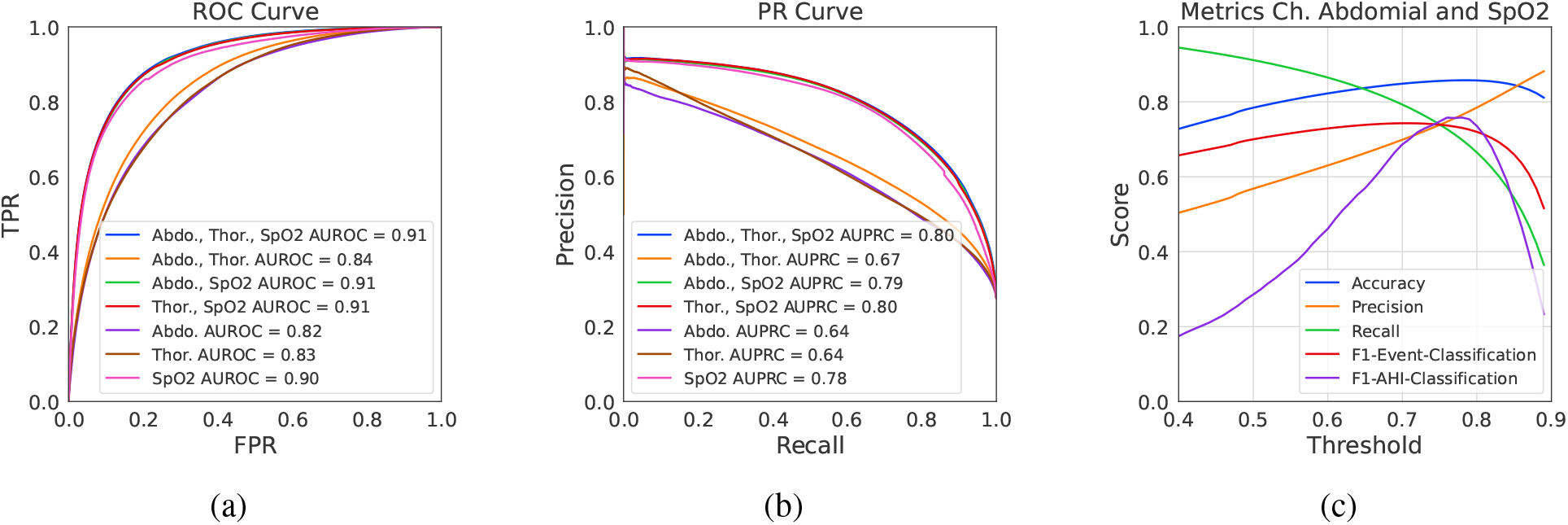
Performance of DRIVEN on classification of AHI-events when ground-truth knowledge on the sleep/awake events is used for all patients in the test dataset. (a) Receiver-operator characteristic and (b) precision-recall curves. (c) Threshold dependent performance metrics for two input channels (abdominal movement and SpO_2_).

**Table S1:** Comparison between different sensor inputs to DRIVEN and its measured AUROC and AUPRC scores for sleep/awake detection on the test and validation datasets in trial 2. The corresponding curves are shown in Figure S1.

| Channels | Test |  | Validation |  |
| --- | --- | --- | --- | --- |
|  | AUROC | AUPRC | AUROC | AUPRC |
| Abdominal + Thoracic + SpO<sub>2</sub> + RRI | 0.900 | 0.924 | 0.893 | 0.958 |
| Abdominal + Thoracic + SpO<sub>2</sub> | 0.901 | 0.925 | 0.894 | 0.959 |
| Abdominal + Thoracic + RRI | 0.887 | 0.915 | 0.888 | 0.957 |
| Abdominal + Thoracic | 0.871 | 0.906 | 0.878 | 0.954 |
| Abdominal + SpO<sub>2</sub> + RRI | 0.878 | 0.907 | 0.859 | 0.944 |
| Abdominal + SpO<sub>2</sub> | 0.873 | 0.903 | 0.854 | 0.942 |
| Thoracic + SpO<sub>2</sub> + RRI | 0.871 | 0.902 | 0.867 | 0.947 |
| Thoracic + SpO<sub>2</sub> | 0.869 | 0.901 | 0.863 | 0.946 |
| Abdominal + RRI | 0.866 | 0.898 | 0.855 | 0.943 |
| Abdominal | 0.850 | 0.890 | 0.842 | 0.940 |
| Thoracic + RRI | 0.857 | 0.892 | 0.864 | 0.947 |
| Thoracic | 0.839 | 0.884 | 0.854 | 0.944 |
| SpO<sub>2</sub> + RRI | 0.758 | 0.797 | 0.730 | 0.870 |
| SpO<sub>2</sub> | 0.716 | 0.758 | 0.678 | 0.841 |
| RRI | 0.702 | 0.751 | 0.705 | 0.860 |

**Table S2:**
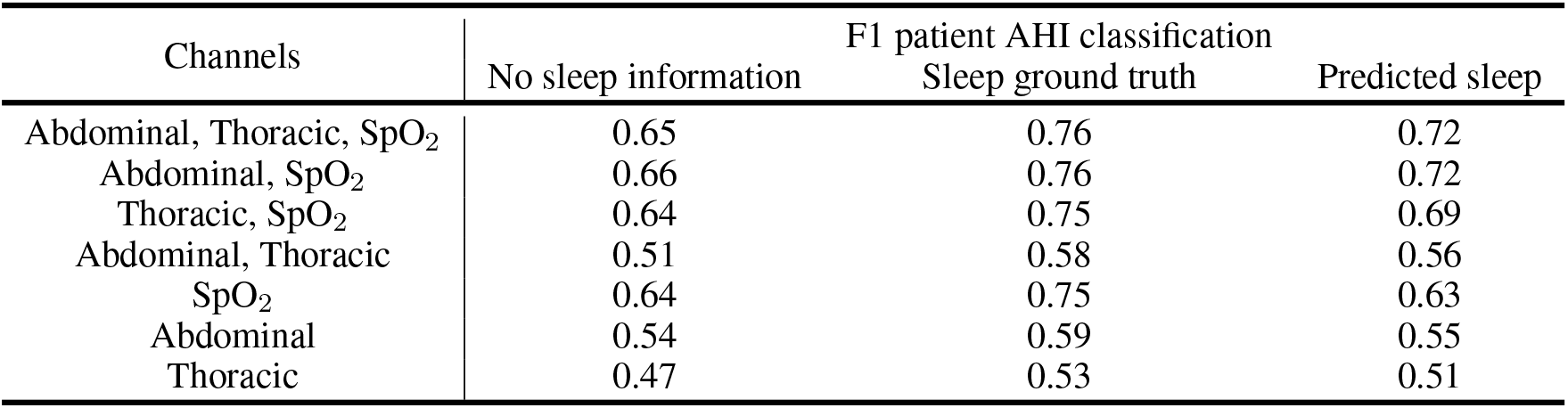
DRIVEN performance for different channel combinations when using no sleep information, corrected with PSG-labeled sleep state, and corrected with DRIVEN’s sleep prediction.

**Table S3:** Comparison of LGBM-3 model performance on AHI-events classification based on different sensor combinations and CNN input window sizes in a 3-fold cross-validation experiment with 1000 randomly selected patients (800 for training, 200 for validation). This is a summary table of the complete experiment provided in Table S4.

| Channels | Window Size |  |  |  |  |  |
| --- | --- | --- | --- | --- | --- | --- |
|  | 10 |  | 30 |  | 80 |  |
|  | AUROC | AUPRC | AUROC | AUPRC | AUROC | AUPRC |
| Abdominal + Thoracic + Airflow + SpO<sub>2</sub> + RRI | 0.899 | 0.593 | 0.921 | 0.719 | 0.923 | 0.793 |
| Abdominal + Thoracic + Airflow + SpO<sub>2</sub> | 0.897 | 0.587 | 0.916 | 0.702 | 0.921 | 0.786 |
| Abdominal + Thoracic + SpO<sub>2</sub> + RRI | 0.889 | 0.563 | 0.914 | 0.697 | 0.920 | 0.786 |
| Abdominal + Thoracic + SpO<sub>2</sub> | 0.888 | 0.561 | 0.913 | 0.690 | 0.914 | 0.770 |
| Abdominal + Airflow + SpO<sub>2</sub> + RRI | 0.890 | 0.578 | 0.911 | 0.693 | 0.916 | 0.777 |
| Abdominal + Airflow + SpO<sub>2</sub> | 0.886 | 0.567 | 0.913 | 0.698 | 0.916 | 0.779 |
| Thoracic + Airflow + SpO<sub>2</sub> + RRI | 0.892 | 0.575 | 0.911 | 0.681 | 0.915 | 0.772 |
| Thoracic + Airflow + SpO<sub>2</sub> | 0.890 | 0.567 | 0.913 | 0.693 | 0.917 | 0.780 |
| Abdominal + Thoracic + Airflow + RRI | 0.846 | 0.464 | 0.886 | 0.635 | 0.899 | 0.738 |
| Abdominal + Thoracic + Airflow | 0.841 | 0.461 | 0.884 | 0.626 | 0.904 | 0.750 |
| Abdominal + SpO<sub>2</sub> + RRI | 0.881 | 0.556 | 0.908 | 0.682 | 0.912 | 0.771 |
| Abdominal + SpO<sub>2</sub> | 0.880 | 0.556 | 0.906 | 0.678 | 0.910 | 0.768 |
| Thoracic + SpO<sub>2</sub> + RRI | 0.878 | 0.535 | 0.906 | 0.669 | 0.902 | 0.735 |
| Thoracic + SpO<sub>2</sub> | 0.878 | 0.532 | 0.903 | 0.659 | 0.909 | 0.751 |
| Airflow + SpO<sub>2</sub> + RRI | 0.877 | 0.551 | 0.901 | 0.668 | 0.903 | 0.755 |
| Airflow + SpO<sub>2</sub> | 0.875 | 0.549 | 0.902 | 0.678 | 0.901 | 0.752 |
| Thoracic + Airflow + RRI | 0.831 | 0.419 | 0.880 | 0.613 | 0.890 | 0.724 |
| Thoracic + Airflow | 0.825 | 0.416 | 0.873 | 0.605 | 0.886 | 0.717 |
| Abdominal + Airflow + RRI | 0.832 | 0.440 | 0.875 | 0.609 | 0.894 | 0.735 |
| Abdominal + Airflow | 0.827 | 0.436 | 0.876 | 0.615 | 0.897 | 0.744 |
| Abdominal + Thoracic + RRI | 0.814 | 0.394 | 0.868 | 0.587 | 0.893 | 0.728 |
| Abdominal + Thoracic | 0.816 | 0.407 | 0.866 | 0.589 | 0.890 | 0.731 |
| SpO<sub>2</sub> + RRI | 0.846 | 0.484 | 0.877 | 0.603 | 0.878 | 0.683 |
| SpO<sub>2</sub> | 0.850 | 0.487 | 0.875 | 0.605 | 0.871 | 0.676 |
| Abdominal + RRI | 0.789 | 0.353 | 0.852 | 0.561 | 0.881 | 0.714 |
| Abdominal | 0.779 | 0.357 | 0.846 | 0.569 | 0.872 | 0.711 |
| Airflow + RRI | 0.785 | 0.346 | 0.836 | 0.543 | 0.850 | 0.660 |
| Airflow | 0.760 | 0.337 | 0.829 | 0.539 | 0.844 | 0.664 |
| Thoracic + RRI | 0.772 | 0.299 | 0.833 | 0.505 | 0.855 | 0.643 |
| Thoracic | 0.769 | 0.296 | 0.834 | 0.517 | 0.857 | 0.659 |
| RRI | 0.576 | 0.134 | 0.621 | 0.230 | 0.663 | 0.326 |

Table S4: Spreadsheet of classification performance on AHI-events classification for validation and test set of different evaluated models for a subset of 1,000 patients. Comparison of the LGBM-3, LGBM-1 and end-to-end CNN models for different sensor combinations (abdominal, thoracic, airflow, SpO_2_ and RRI) and CNN input window sizes (10, 30 and 80 seconds) in a 3-fold cross-validation experiment with 1000 randomly selected patients (800 for training, 200 for validation). This table is included as a separate spreadsheet file.

Table S5: Spreadsheet of classification performance on AHI-events classification and AHI estimation for validation and test set for three complete data splits, with pre selected models from Table S4. Comparison of the LGBM-3 and LGBM-1 different sensor combinations (abdominal, thoracic, airflow and SpO_2_) and CNN input window sizes (30 and 80 seconds). This table is included as a separate spreadsheet file.

## References

1. Senaratna, C. V. et al. Prevalence of obstructive sleep apnea in the general population: a systematic review. Sleep Medicine Reviews 34, 70–81 (2017).

2. Riha, R. L. Defining obstructive sleep apnoea syndrome: a failure of semantic rules. Breathe 17, 210082 (2021).

3. Mannarino, M. R., Di Filippo, F. & Pirro, M. Obstructive sleep apnea syndrome. European Journal of Internal Medicine 23, 586–593 (2012).

4. Punjabi, N. M. The epidemiology of adult obstructive sleep apnea. Proceedings of the American Thoracic Society 5, 136–143 (2008).

5. McNicholas, W. T. Diagnosis of obstructive sleep apnea in adults. Proceedings of the American Thoracic Society 5, 154–160 (2008).

6. Berry, R. B. et al. Rules for scoring respiratory events in sleep: update of the 2007 AASM manual for the scoring of sleep and associated events: deliberations of the sleep apnea definitions task force of the American Academy of Sleep Medicine. Journal of Clinical Sleep Medicine 8, 597–619 (2012).

7. Hutchison, K. N., Song, Y., Wang, L. & Malow, B. A. Analysis of sleep parameters in patients with obstructive sleep apnea studied in a hospital vs. a hotel-based sleep center. Journal of Clinical Sleep Medicine 4, 119–122 (2008).

8. Levy, J., Álvarez, D., Del Campo, F. & Behar, J. A. Deep learning for obstructive sleep apnea diagnosis based on single channel oximetry. Nature Communications 14, 4881 (2023).

9. Ramachandran, A. & Karuppiah, A. A survey on recent advances in machine learning based sleep apnea detection systems. Healthcare 9, 914 (2021).

10. Mendonca, F., Mostafa, S. S., Ravelo-Garcia, A. G., Morgado-Dias, F. & Penzel, T. A review of obstructive sleep apnea detection approaches. IEEE Journal of Biomedical and Health Informatics 23, 825–837 (2018).

11. Mostafa, S. S., Mendonça, F. G. Ravelo-García, A. & Morgado-Dias, F. A systematic review of detecting sleep apnea using deep learning. Sensors 19, 4934 (2019).

12. Yumino, D. et al. Differing effects of obstructive and central sleep apneas on stroke volume in patients with heart failure. American Journal of Respiratory and Critical Care Medicine 187, 433–438 (2013).

13. Faust, O., Hagiwara, Y., Hong, T. J., Lih, O. S. & Acharya, U. R. Deep learning for healthcare applications based on physiological signals: A review. Computer Methods and Programs in Biomedicine 161, 1–13 (2018).

14. Tang, J., Alelyani, S. & Liu, H. Feature selection for classification: A review. Data Classification: Algorithms and Applications, 37 (2014).

15. Ganapathy, N., Swaminathan, R. & Deserno, T. M. Deep learning on 1-D biosignals: a taxonomy-based survey. Yearbook of Medical Informatics 27, 098–109 (2018).

16. Chen, X. et al. Racial/ethnic differences in sleep disturbances: the Multi-Ethnic Study of Atherosclerosis (MESA). Sleep 38, 877–888 (2015).

17. Blackwell, T. et al. Associations between sleep architecture and sleep-disordered breathing and cognition in older community-dwelling men: the osteoporotic fractures in men sleep study. Journal of the American Geriatrics Society 59, 2217–2225 (2011).

18. Quan, S. F. et al. The sleep heart health study: design, rationale, and methods. Sleep 20, 1077–1085 (1997).

19. Zhang, G.-Q. et al. The National Sleep Research Resource: towards a sleep data commons. Journal of the American Medical Informatics Association 25, 1351–1358 (2018).

20. Arnold, J., Boucher, J., Mobley, D., Nawabit, R. & Redline, S. SRC Manual of Operations and Scoring Rules (MESA PSG Sleep Reading Center, 2014).

21. Ke, G. et al. Lightgbm: A highly efficient gradient boosting decision tree. Advances in Neural Information Processing Systems 30 (2017).

22. Mohammed, R., Rawashdeh, J. & Abdullah, M. Machine learning with oversampling and undersampling techniques: overview study and experimental results. 11th International Conference on Information and Communication Systems, 243–248 (2020).

23. Bentéjac, C., Csörgő, A. & Martínez-Muñoz, G. A comparative analysis of gradient boosting algorithms. Artificial Intelligence Review 54, 1937–1967 (2021).

24. LeCun, Y., Bengio, Y. & Hinton, G. Deep learning. en. Nature 521, 436–444. ISSN: 0028-0836, 1476-4687 (May 2015).

25. Sutton, R. The bitter lesson. Incomplete Ideas (blog) 13, 38 (2019).

26. Moridian, P. et al. Automatic diagnosis of sleep apnea from biomedical signals using artificial intelligence techniques: Methods, challenges, and future works. WIREs Data Mining and Knowledge Discovery 12 (2022).

27. Chen, S., Redline, S., Eden, U. T. & Prerau, M. J. Dynamic models of obstructive sleep apnea provide robust prediction of respiratory event timing and a statistical framework for phenotype exploration. Sleep 45, zsac189 (2022).

28. Gu, A. & Dao, T. Mamba: Linear-Time Sequence Modeling with Selective State Spaces. 2312.00752 (2023).

29. Alvarez-Estevez, D. & Moret-Bonillo, V. Computer-assisted diagnosis of the sleep apnea-hypopnea syndrome: a review. Sleep Disorders 2015, 237878 (2015).

30. Kemp, B. & Olivan, J. European data format ‘plus’(EDF+), an EDF alike standard format for the exchange of physiological data. Clinical Neurophysiology 114, 1755–1761 (2003).

31. Rechtschaffen, A. A manual for standardized terminology, techniques and scoring system for sleep stages in human subjects. Brain Information Service 20, 246–247 (1969).

32. Huang, L. et al. Normalization techniques in training DNNs: Methodology, analysis and application. arxiv:2009.12836 (2020).

33. Pan, J. & Tompkins, W. J. A real-time QRS detection algorithm. IEEE Transactions on Biomedical Engineering BME-32, 230–236 (1985).

34. Sedghamiz, H. Matlab implementation of Pan Tompkins ECG QRS detector. Code available at the File Exchange Site of Mathworks. https://www.mathworks.com/matlabcentral/fileexchange/45840-complete-pan-tompkins-implementation-ecg-qrs-detector (2014).

35. Keys, R. Cubic convolution interpolation for digital image processing. IEEE Transactions on Acoustics, Speech, and Signal Processing 29, 1153–1160 (1981).

36. Tan, M. & Le, Q. Efficientnetv2: Smaller models and faster training. International Conference on Machine Learning, 10096–10106 (2021).

37. Tan, M. & Le, Q. Efficientnet: Rethinking model scaling for convolutional neural networks. International Conference on Machine Learning, 6105–6114 (2019).

38. He, K., Girshick, R. & Dollár, P. Rethinking imagenet pre-training. IEEE/CVF International Conference on Computer Vision, 4918–4927 (2019).

39. Jais, I. K. M., Ismail, A. R. & Nisa, S. Q. Adam optimization algorithm for wide and deep neural network. Knowledge Engineering and Data Science 2, 41–46 (2019).

40. Chen, T. et al. Xgboost: extreme gradient boosting. R Package Version 0.4-2 1, 1–4 (2015).

41. Ye, J., Chow, J.-H., Chen, J. & Zheng, Z. Stochastic gradient boosted distributed decision trees. ACM Conference on Information and Knowledge Management, 2061–2064 (2009).

42. Bergstra, J. & Bengio, Y. Random search for hyper-parameter optimization. Journal of Machine Learning Research 13 (2012).

43. Kotzen, K. et al. SleepPPG-Net: A Deep Learning Algorithm for Robust Sleep Staging From Continuous Photoplethysmography. IEEE Journal of Biomedical and Health Informatics 27, 924–932 (2023).

44. Varrette, S., Bouvry, P., Cartiaux, H. & Georgatos, F. Management of an Academic HPC Cluster: The UL Experience. International Conference on High Performance Computing & Simulation, 959–967 (2014).

